# Mapping Evidence on Sports-Related Thumb Injuries: Scoping Review Protocol

**DOI:** 10.1101/2025.10.20.25338370

**Authors:** Valentina Scalise, Niccolò D’Agostino, Sara Di Serio, Silvia Minnucci

**Affiliations:** Department of Clinical Science and Translational Medicine, University of Rome Tor Vergata, Rome, Italy

**Keywords:** Sport Injuries, Thumb, Ulnar Collateral Ligament, De Quervain, Athletes, Ligament Injuries, Fractures, Scoping Review

## Abstract

**Background:** “Thumb injuries represent a significant concern in the context of sports, substantially affecting hand function, athletic performance, and return to sport. Despite the high incidence of conditions such as fractures, ligament injuries (e.g., ulnar collateral ligament tears), post-traumatic tenosynovitis (e.g., extensor pollicis longus [EPL] tendinopathy), joint capsule injuries (e.g., metacarpophalangeal dislocations), and De Quervain’s tenosynovitis, the scientific literature lacks a comprehensive synthesis of the available evidence.”

**Objective:** To provide a comprehensive overview of studies addressing both traumatic and overuse-related thumb injuries in athletes, synthesizing data by sport type, injury mechanism, diagnosis, treatment (conservative or surgical), and return to sport outcomes. Additionally, gaps in existing literature and future research priorities will also be identified. An infographic will be developed to support clinicians, physical therapists, and athletes by translating the main findings into practical information regarding injury mechanisms, treatment strategies, and return-to-sport considerations.

**Methods:** This scoping review will be conducted in accordance with the methodological framework outlined in the JBI Manual for Evidence Synthesis. The databases PubMed, Scopus, CINAHL, Cochrane Library, and PEDro will be searched for studies published from 2015 onwards, in English or Italian. In addition, Google Scholar will be consulted to identify relevant grey literature. Studies from any setting and involving athletes of all levels will be eligible for inclusion. One reviewer will screen all records and perform full data extraction. Two reviewers will independently check a randomly assigned subset of records to ensure reliability. A fourth reviewer will act as arbitrator and resolve any conflicts between reviewer 1, 2, and 3. Results will be illustrated using descriptive statistics and summarized in an infographic to facilitate clinical applicability.

**Ethics and Dissemination:** This scoping review does not require ethics approval, as it involves the analysis of previously published data and does not include human participants. An infographic summarizing the main findings will be developed and shared with healthcare professionals, including physical therapists, and with athletes, to provide practical information on injury mechanisms, treatment strategies, and return-to-sport considerations related to thumb injuries. The results will be submitted for publication in peer-reviewed journals and presented at relevant national and international conferences. Based on the findings, recommendations will be formulated to guide future research directions in this field.

## INTRODUCTION

Thumb injuries are common in sports, particularly in disciplines involving high-impact forces, grip, stress, or falls.These injuries can substantially impair hand function, negatively affect athletic performance, and delay return to sport activities [5,7,8,10].The reported incidence of thumb injuries in athletes varies across disciplines and sports modalities. In alpine skiing, for example, up to 16–25% of all upper limb injuries involve the thumb, predominantly due to UCL lesions [10]. In ball-handling sports like handball and basketball, thumb injuries account for approximately 10– 20% of all hand and wrist injuries, with ligamentous tears being among the most common [5,10]. Although the overall prevalence across all sports is less clearly defined, thumb pathologies represent a significant portion of upper extremity injuries in elite and recreational athletes [10].

The clinical spectrum of thumb injuries is broad, encompassing both traumatic and non-traumatic entities that may considerably affect athletic populations.Traumatic injuries include dislocations— mainly at the metacarpophalangeal and interphalangeal joints—as well as fractures, which may involve the distal phalanx, proximal phalanx, first metacarpal, or present as Bennett and Rolando fractures.These injuries may involve ligamentous structures ulnar collateral ligament [UCL] or radial collateral ligament [RCL]), and tendons (e.g., De Quervain’s tenosynovitis), and their mechanisms are often sport-specific [5,7]. Rare but complex injuries like combined RCL and UCL ruptures in adolescents have also been reported [11].

The skier’s thumb, a frequent UCL injury due to forced abduction of the thumb, is well-documented in winter sports [3], while ball-handling sports like basketball and handball often report collateral ligament ruptures due to valgus or varus stress [4,7]. Although most studies report on traumatic injuries, chronic conditions such as tendinopathies can occur with overuse, especially in repetitive motion sports [6].

Proper diagnosis and treatment of these conditions are crucial, especially in the context of return-to-sport decisions [9]. Despite the prevalence of these injuries, no comprehensive mapping exists summarizing the full spectrum of thumb pathologies in athletes, including both traumatic and non-traumatic entities.

The aim of this scoping review is therefore to map the range of thumb injuries affecting athletes, detailing their nature, diagnostic pathways, and outcomes, because a comprehensive synthesis of the literature summarizing the full spectrum of sport-related thumb pathologies—both traumatic and non-traumatic—is lacking. [8].

### OBJECTIVES

- Identify, classify, and describe the principal traumatic, non-traumatic and overuse-related thumb injuries in athletes, including their underlying mechanisms, diagnostic approaches, and therapeutic strategies.
- Summarize studies according to sport type, patient characteristics (e.g., age, competition level), and specific injury type/problematic.
- Explore outcomes related to return to sport
- To identify potential gaps in the existing literature and suggest areas for future research.
- Inform clinicians and patients through the realization of an infographic that summarizes the main findings, providing practical insights for clinical decision-making and injury prevention.

## METHODS

This scoping review was conducted according to the Joanna Briggs Institute (JBI) methodology for scoping reviews [1] and adheres to the PRISMA Extension for Scoping Reviews (PRISMA-ScR) reporting guidelines [2].

The 6-stage framework proposed by Arksey and O’Malley was followed [13]:

(1)identification of the research question; (2) identification of relevant studies; (3) selection of studies; (4) charting of data; (5) summary and reporting of results; and (6) an optional consultation exercise, which was not conducted in this study as it was deemed unnecessary.

The PCC framework (Population, Concept, Context) recommended by JBI was used to define the inclusion criteria. The protocol structure also follows the model proposed by Lely et al. [11].

This scoping review protocol was registered in medRxiv.

### Review questions

This scoping review aims to answer the following research questions:

1. Which types of traumatic and non-traumatic thumb injuries are reported in sport participants (recreational, amateur/collegiate, and professional)?
2. In which sports are thumb injuries most frequently reported?
3. What are the mechanisms most commonly associated with these injuries?
4. Which diagnostic methods and therapeutic interventions are used to manage thumb injuries in sport participants (acute vs. overuse)?
5. What functional and clinical outcomes are reported following treatment of sport-related thumb injuries?
6. What return-to-sport outcomes are described, including time to return, rate of return at the same or lower level, and factors influencing successful return?

### Eligibility Criteria

This scoping review will consider studies of any design and publication type to ensure a comprehensive overview of thumb injuries in sport participants. However, primary studies that have already been incorporated within existing evidence syntheses or systematic reviews will generally be excluded, unless they contain unique data not otherwise reported in those syntheses. Research conducted on animals and in vitro studies will be excluded, articles not published in English or Italian will be excluded. Importantly, to capture the most recent and relevant evidence, only studies published within the 10 years preceding the date of the search will be included (25 August 2015 – 25 August 2025). This temporal limit is intended to reflect current understandings and advances in diagnosis, treatment, and outcomes related to sports-related thumb injuries. Detailed inclusion and exclusion criteria are summarized in Table I.

**Table I.**

|  |  |
| --- | --- |
| **Population** | This review will include studies involving sport participants of any age, sex, or competitive level, encompassing recreational, amateur, semi-professional, and professional participants. Both male and female |
|  | participants in individual sports (e.g., skiing, golf, racquet sports, bowling) as well as team sports (e.g., basketball, handball, baseball, football) will be considered. Age groups include children, adolescents, and adults, to capture variation in injury patterns across developmental stages. The inclusion of diverse sports and athlete profiles ensures a comprehensive epidemiological and clinical overview of thumb injuries in all relevant sport populations. |
| **Concept** | This review includes traumatic and overuse-related thumb injuries, including fractures (e.g., phalangeal, metacarpal, Bennett, Rolando), ligament injuries (ulnar collateral ligament — UCL, radial collateral ligament — RCL tears), dislocations, tendinopathies (e.g., De Quervain's tenosynovitis, extensor pollicis longus - EPL tendinopathy), and post-traumatic conditions (e.g., instability, synovitis, degenerative changes). Both acute and chronic conditions are included. The review also considers less common or rare entities such as spontaneous tendon ruptures or vascular complications related to thumb trauma in sport. Diagnostic approaches, treatment modalities (conservative and surgical), and return-to-sport outcomes are included as well, reflecting the clinical complexity of thumb injuries in sport participants.[5,6,7] |
| **Context** | Any sport-related setting will be included, with no restrictions on geographical location, type or level of sport, or care setting. |
| **Table I.** |  |

## Information Sources and Search Strategy

The search strategy was developed by four reviewers to ensure a rigorous and comprehensive overview of the literature on thumb injuries in sport participants populations. The research team included authors with expertise in physiotherapy, sports injury management, and evidence synthesis methodology.

A preliminary search was conducted to identify relevant terms and refine the final search strategy. The databases searched included MEDLINE central, Scopus, CINAHL, Cochrane Central, and PEDro which were selected due to their extensive coverage of biomedical and sports medicine literature. In addition, the reference list of included studies and at least the first 10-12 pages of Google Scholar were searched.

The search was restricted to studies published in English or Italian language within the last 10 years ((“2015/01/01”[PDAT] : “2025/08/25”[PDAT]), and was conducted in August 2025 in, Carpino(FG), Italy. Boolean operators, Medical Subject Headings (MeSH), and free-text terms were employed to maximize the identification of relevant articles, and the search strategy was peer-reviewed where possible (PRESS).

In addition to database searches, the reference lists of included studies were manually screened for additional eligible articles, and selected gray literature sources (conference abstracts, trial registries) were searched when feasible. The search strategy is reported in Table I and Table II. The PRISMA-S guidelines were followed for reporting the search strategy. Search results were imported into Rayyan, a web-based tool that facilitated duplicate removal and screening[14].

**Table II.**

|  |  |
| --- | --- |
| **Population (P)** | "Athletes"[MeSH], "Athletic Injuries"[MeSH], "Sports"[MeSH], Free-text: athlete*, sport*, player*, elite, amateur*, professional*, "sport participant" |
| **Concept (C)** | "Thumb"[MeSH], "Thumb Injuries"[MeSH], "Metacarpal Bones"[MeSH], "Finger Phalanges"[MeSH], "Carpometacarpal Joints"[MeSH], "Metacarpophalangeal Joint"[MeSH], "Collateral Ligaments"[MeSH], "Fractures, Bone"[MeSH], "Ligament Injuries"[MeSH], "Tendinopathy"[MeSH], "Tenosynovitis"[MeSH], "De Quervain Disease"[MeSH], "Pain"[MeSH]<br>Free-text: "thumb injury", "thumb injuries", "thumb fracture", "thumb trauma", "skier's thumb", "gamekeeper's thumb", "ulnar collateral ligament", UCL, "radial collateral ligament", RCL, Stener, "Bennett fracture", "Rolando fracture", "thumb dislocation", "De Quervain", tendinopath*, tenosynoviti* |
| **Context (C)** | "Sports"[MeSH], "Sports Medicine"[MeSH]<br>Free-text: sport*, athlete*, player*, elite, amateur*, professional* |

**Table III.**
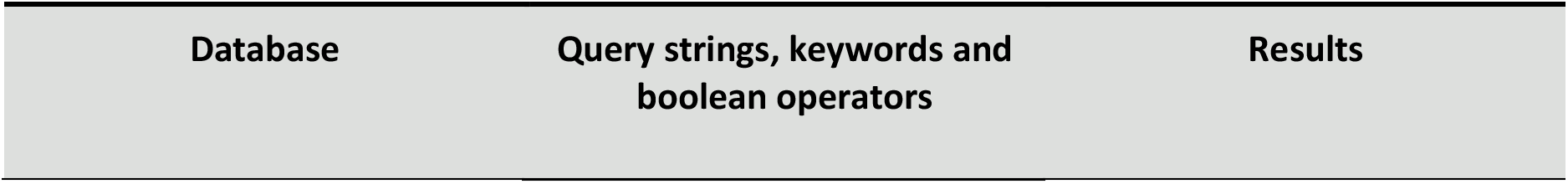
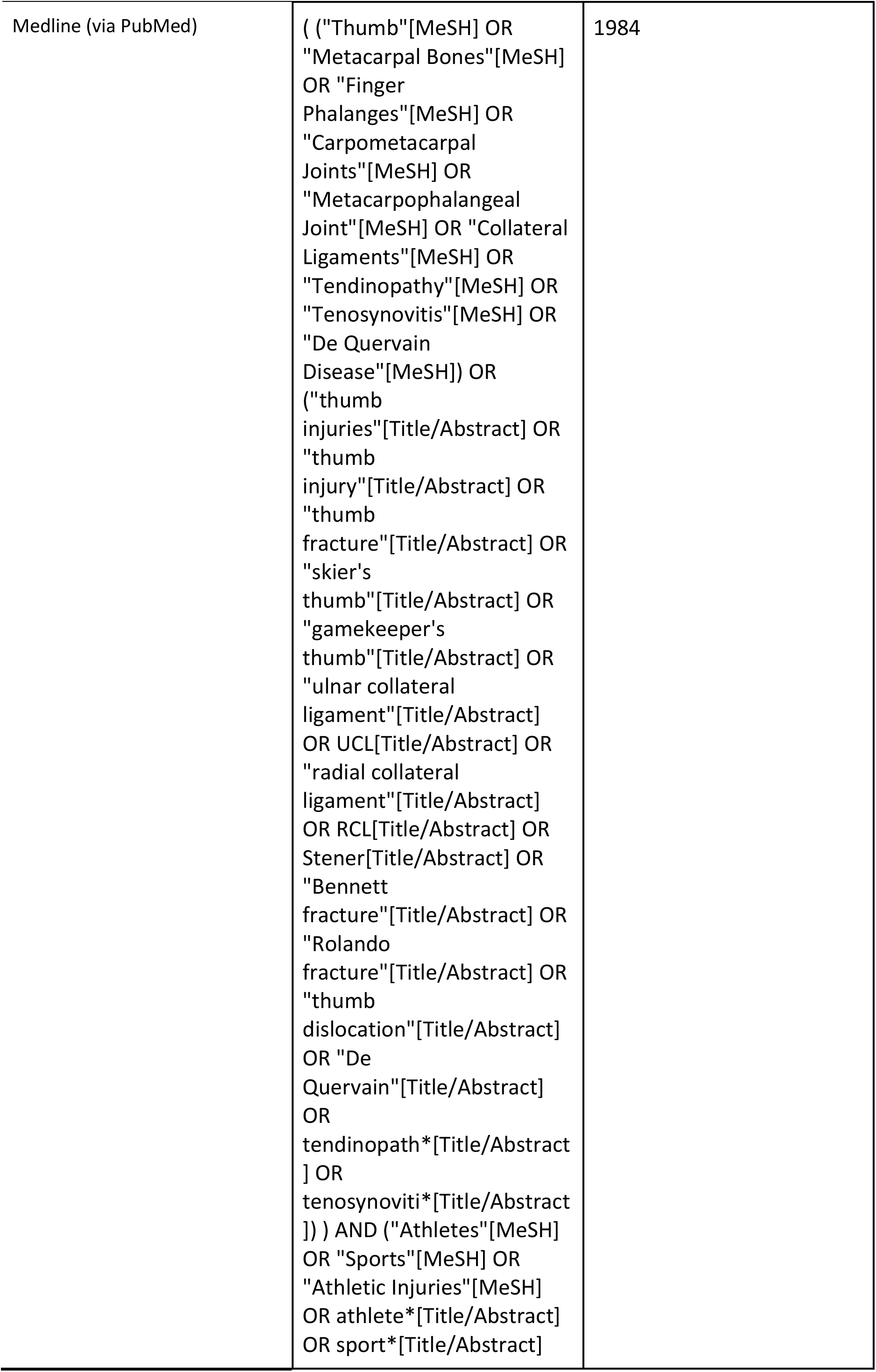
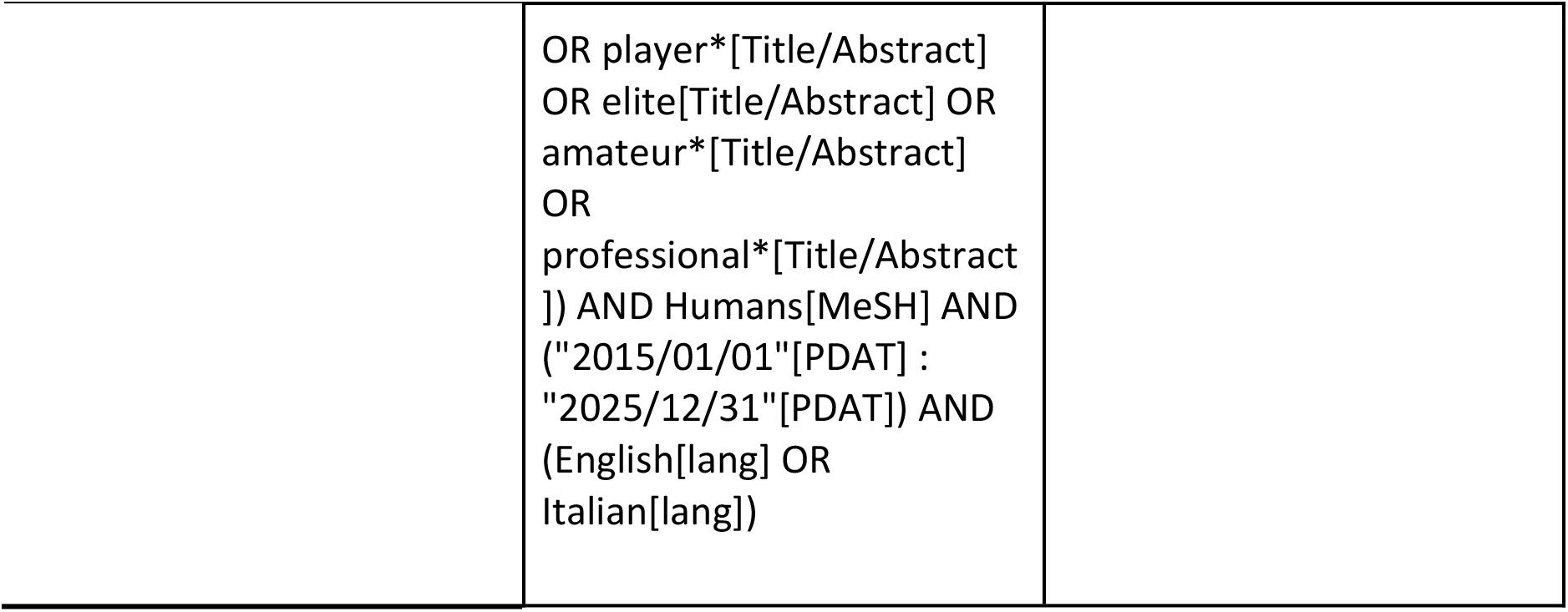
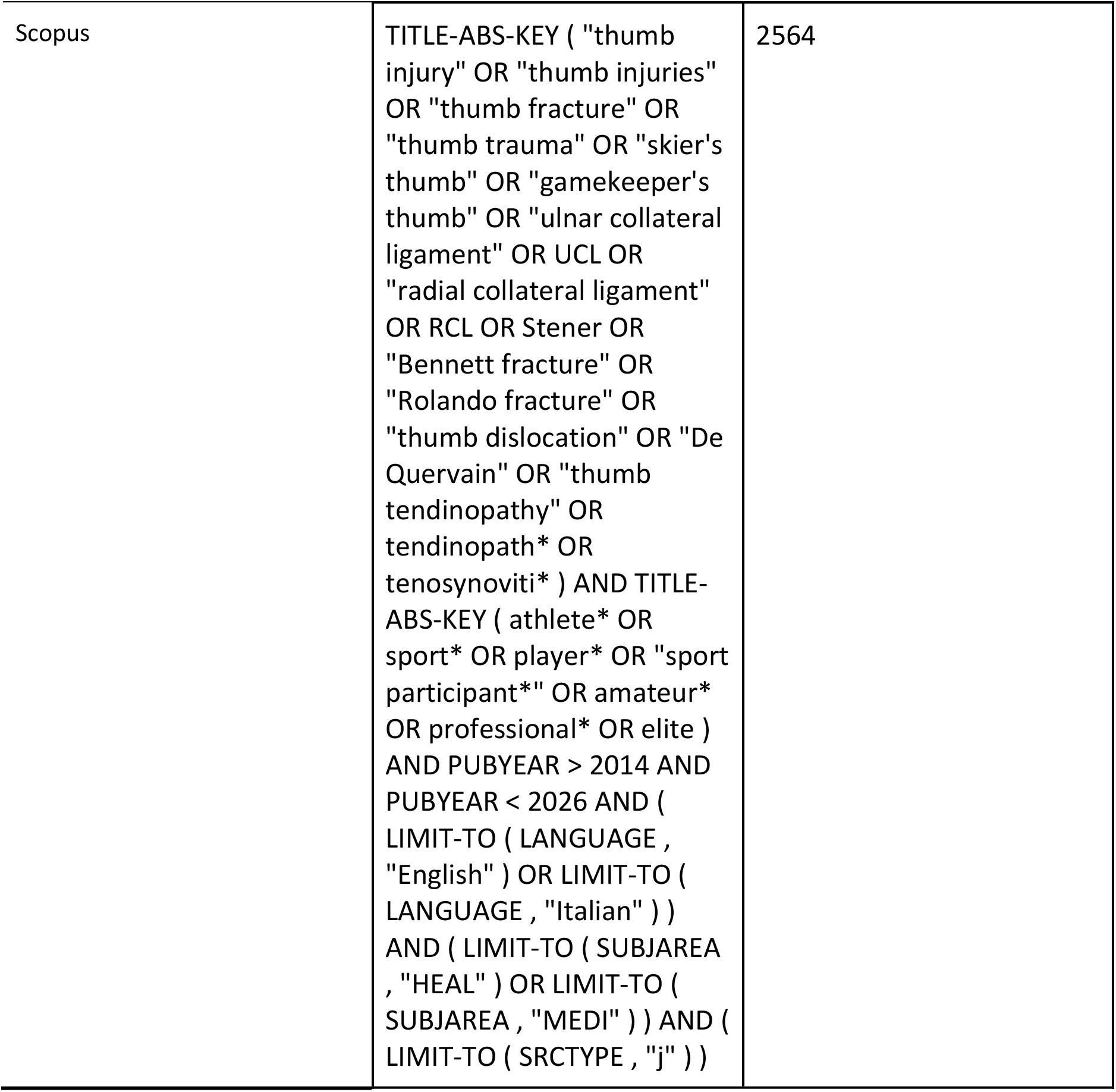
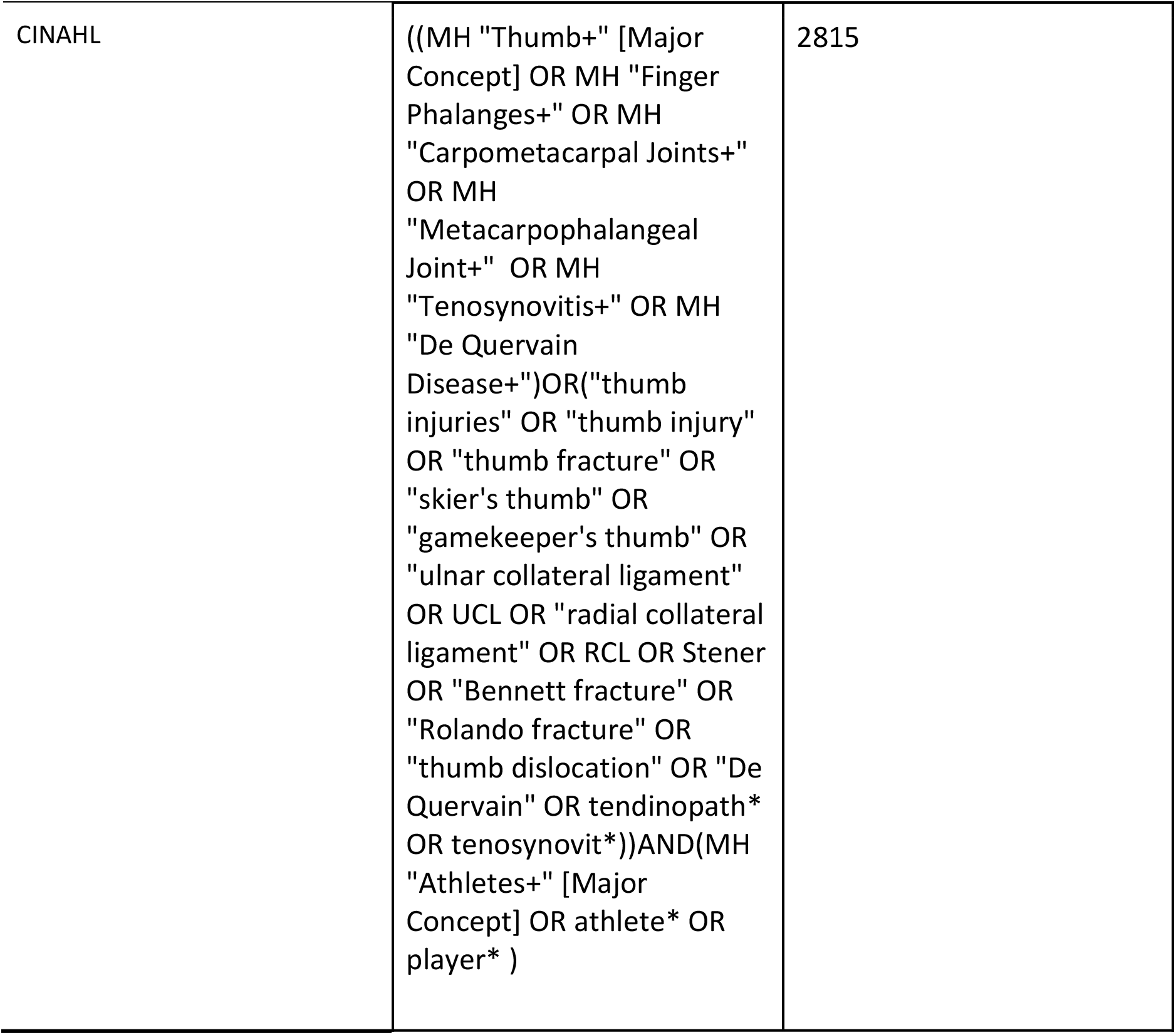
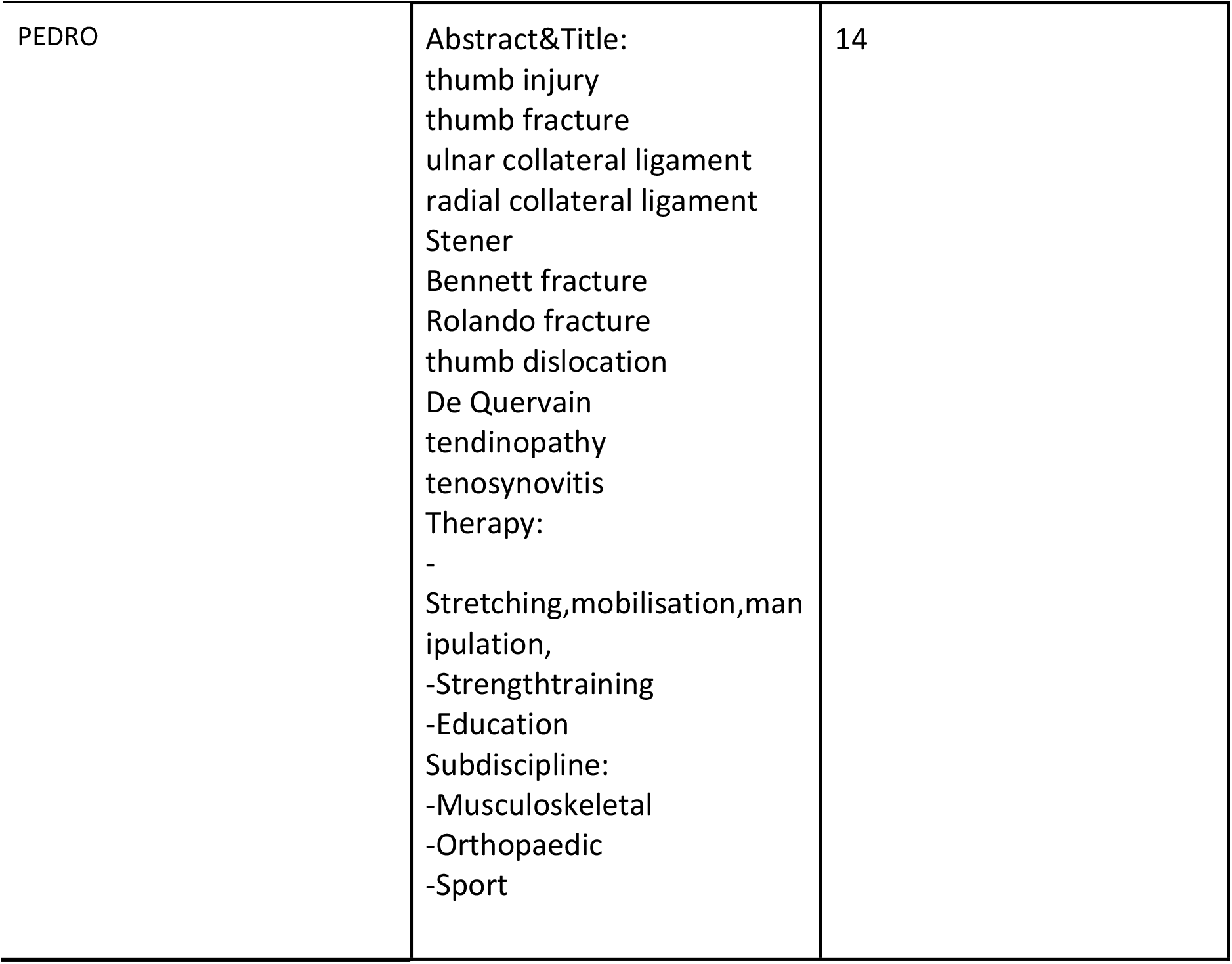
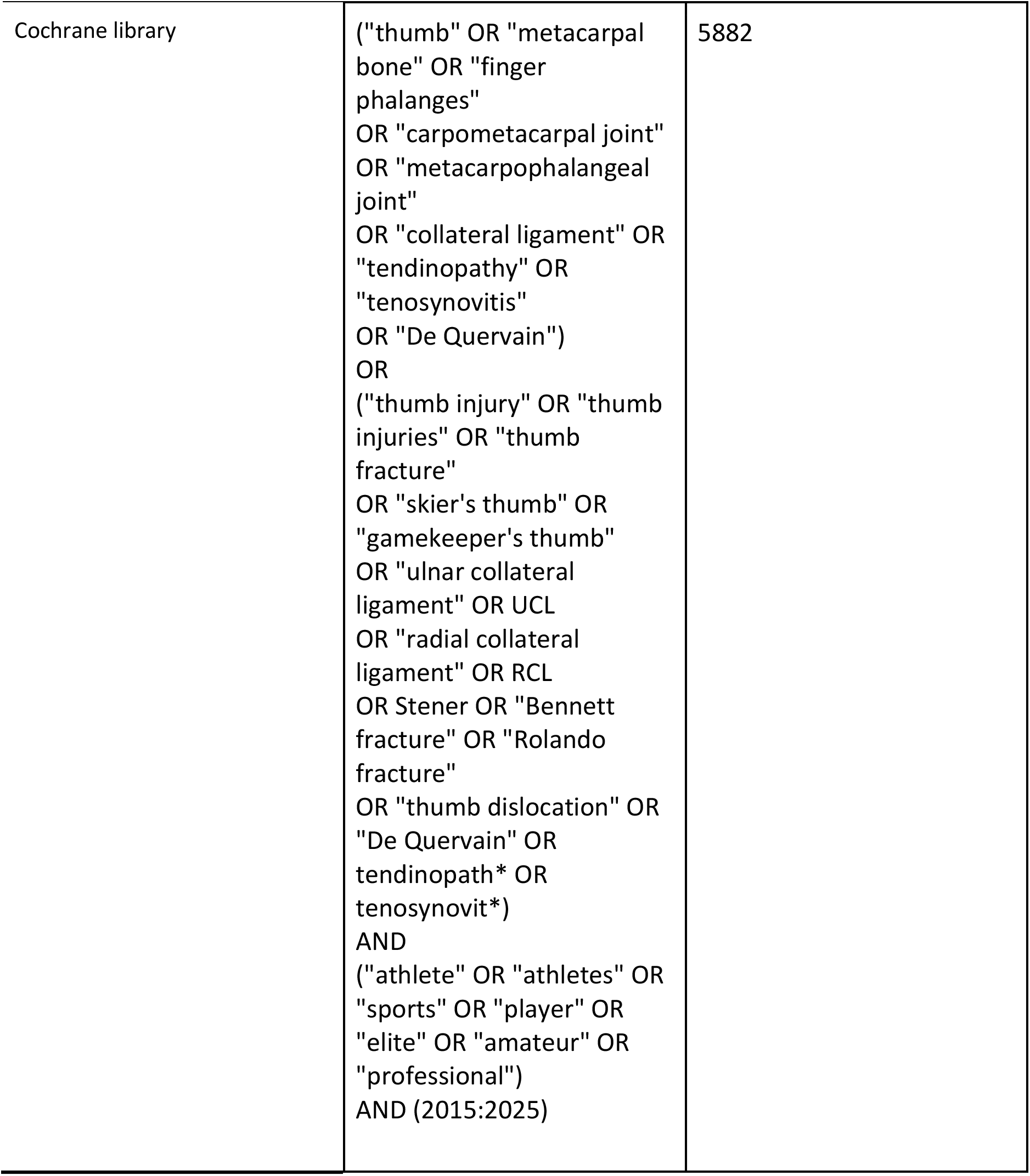
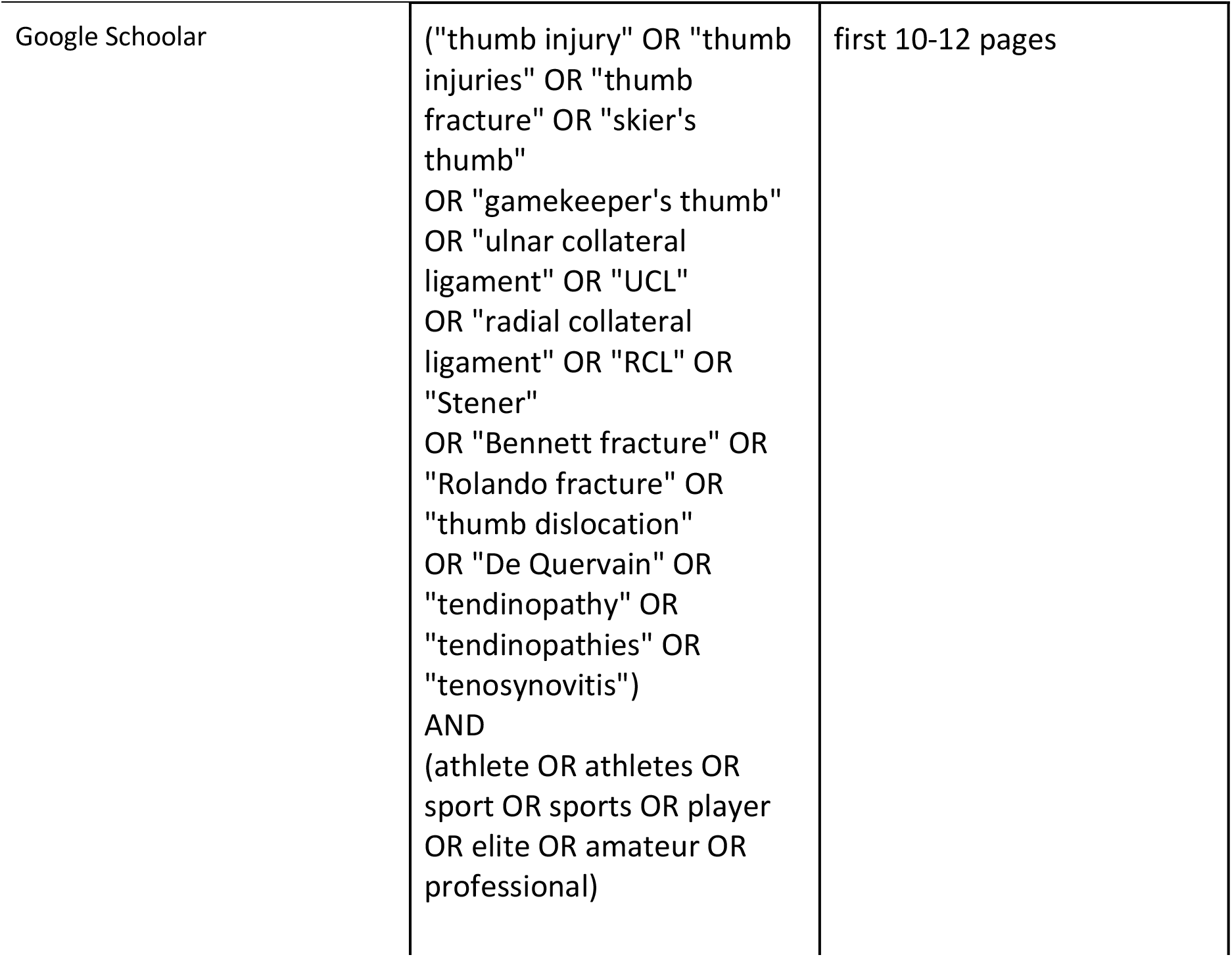

### Inclusion Criteria

- Original research (quantitative or qualitative), review articles, case reports or case series, and relevant grey literature (e.g., theses, institutional reports, conference proceedings, preprints).
- Articles published in English or Italian between January 1, 2015, and August 25, 2025.
- Studies involving sport participants at any competitive level, including recreational, amateur, collegiate, professional, or elite athletes.

### Exclusion Criteria

- Studies not specific to the thumb
- Studies not involving sport participants
- Publications in languages other than English or Italian

## STUDY SELECTION AND DATA EXTRACTION

The web-based platform Rayyan (Rayyan Systems Incorporation, Cambridge, USA) was used for the entire selection process, including duplicate removal and screening. The Cochrane Handbook for Systematic Reviews of Interventions (version 6.4, updated August 2023) was consulted for methodological guidance on study selection and data management [15].The screening process was conducted independently by three reviewers. One reviewer screened all records and performed full data extraction, while two reviewers independently assessed a randomly assigned subset of records to ensure reliability. A fourth reviewer resolved any conflicts between reviewer 1, 2, and 3.. Articles without an abstract were automatically moved to full-text screening. When full-texts were not accessible, study authors were contacted up to two times (one attempt per week). Throughout the process, the review team met regularly to clarify objectives, address discrepancies, and monitor progress according to predefined timelines. A detailed list of excluded studies with the reasons for exclusion is reported in Appendix II.

Data extraction was also performed independently by three reviewers, following the same approach used for study selection. Relevant information was charted in a Microsoft Excel spreadsheet (Microsoft Corporation, Redmond, WA, USA), which is available in the appendix. Extracted items included:

1. Study characteristics: design, authors, year of publication
2. Aims of the study
3. Population details: sample size, demographics (age, sex), sport discipline, and level of participation (recreational, amateur, professional)
4. Type of thumb injury: traumatic (fractures, ligament injuries, dislocations) or non-traumatic/overuse (tendinopathies, De Quervain’s, post-traumatic instability, degenerative changes)
5. Diagnostic methods: clinical evaluation, imaging techniques, functional tests
6. Treatment strategies: conservative (rehabilitation, splinting, physical therapy) and surgical (repair, reconstruction), as well as feasibility, safety, and accessibility
7. Context: sport environment, competition level, and geographical setting
8. Outcomes: especially return-to-sport (time, rate), functional recovery, complications, and other reported clinics.

## Data Availability

All data produced in the present work are contained in the manuscript

## ETHICS AND DISSEMINATION

This scoping review does not require ethics approval as it involves the synthesis of already published literature. An infographic summarizing the main findings will be developed to support dissemination among clinicians, athletes, and physical therapists, facilitating evidence-informed practice and patient education. The results may also inform recommendations for future research in the field of thumb injuries in athletes.

## CONTRIBUTORS

The review project was initiated and developed by the author as part of an academic thesis. The review project was initiated by Valentina Scalise, who also defined the framework. The author conceived the research framework, designed the protocol, conducted the search, screening, and data extraction processes, prepared the manuscript and approved the final version.

## ACKNOWLEDGEMENT

Nothing to report.

## References

1. Peters MDJ, Godfrey CM, McInerney P, et al. Chapter 11: Scoping Reviews. In: JBI Manual for Evidence Synthesis. JBI, 2020. doi:10.46658/JBIMES-20-12

2. Tricco AC, Lillie E, Zarin W, et al. PRISMA Extension for Scoping Reviews (PRISMA-ScR): Checklist and Explanation. Ann Intern Med. 2018;169(7):467–467. doi:10.7326/M18-0850

3. Mahajan M, Tolman C, Würth B, Rhemrev SJ. Clinical evaluation vs magnetic resonance imaging of the skier’s thumb: A prospective cohort of 30 patients. Eur J Radiol. 2016;85(10):1750–1750. doi:10.1016/j.ejrad.2016.07.007

4. Avery DM 3rd, Caggiano NM, Matullo KS. Ulnar collateral ligament injuries of the thumb: a comprehensive review. Orthop Clin North Am. 2015 Apr;46(2):281–292. doi:10.1016/j.ocl.2014.11.007

5. Kadow TR, Fowler JR. Thumb Injuries in Athletes. *Hand Clin.* 2017 Feb;33(1):161–173. doi:10.1016/j.hcl.2016.08.008

6. Ilyas AM, Ast M, Schaffer AA, Thoder J. De Quervain tenosynovitis of the wrist. J Am Acad Orthop Surg. 2007;15(12):757–757. doi:10.5435/00124635-200712000-00006

7. Robinson DM, Kakar S, Jelsing E. Acute Thumb Metacarpophalangeal Joint Ulnar Collateral Ligament Injury: Diagnosis, Management, and Return to Sports Considerations. Curr Sports Med Rep. 2023;22(6):238–238. doi:10.1249/JSR.0000000000001079

8. Ryan L, Doody O. The treatment, outcomes and management of hand, wrist, finger, and thumb injuries in the professional/amateur contact sport athletes: A scoping review. Int J Orthop Trauma Nurs. 2024; doi:10.1016/j.ijotn.2024.101108

9. Allahabadi S, Kwong JW, Pandya NK, et al. Return to Play After Thumb Ulnar Collateral Ligament Injuries Managed Surgically in Athletes—A Systematic Review. J Hand Surg Glob Online. 2023;5(4):234–234. doi:10.1016/j.jhsg.2023.03.005

10. Lehman JD, Krishnan KR, Stepan JG, Nwachukwu BU. Prevalence and Treatment Outcomes of Hand and Wrist Injuries in Professional Athletes: A Systematic Review. HSS J. 2020;16(4):336–336. doi:10.1007/s11420-020-09760-w

11. Bhat AK, Mane PP, Acharya A, Madi S. Simultaneous combined complete tear of radial and ulnar collateral ligaments of thumb in an adolescent. BMJ Case Rep. 2017;2017:bcr-2017-220550. doi:10.1136/bcr-2017-220550

12. Lely J, Morris HC, Sasson N, et al. How to write a scoping review protocol: Guidance and template. Published online May 30, 2023. doi:10.17605/OSF.IO/YM65X

13. Arksey H, O’Malley L. Scoping studies: towards a methodological framework. Int J Soc Res Methodol. 2005;8(1):19–32. doi:10.1080/1364557032000119616

14. Ouzzani M, Hammady H, Fedorowicz Z, Elmagarmid A. Rayyan—a web and mobile app for systematic reviews. Syst Rev. 2016;5(1):210–210. doi:10.1186/s13643-016-0384-4

15. Higgins JPT, Thomas J, Chandler J, et al., editors. Cochrane Handbook for Systematic Reviews of Interventions, version 6.4 (updated August 2023). Cochrane; 2023. Available from: https://training.cochrane.org/handbook

